# Polygenic Susceptibility to Diabetes and Poor Glycemic Control in Stroke Survivors

**DOI:** 10.1101/2023.09.18.23295736

**Authors:** Zachariah S. Demarais, Carolyn J. Conlon, Shufan Huo, Cyprien Rivier, Daniela Renedo, Kevin N. Sheth, Guido J. Falcone

## Abstract

**Importance:** Type 2 diabetes mellitus is a highly heritable disease with numerous identified genetic risk variants. However, the concrete role of these variants in the clinical care of stroke patients remains poorly understood.

**Objective:** To evaluate whether higher polygenic susceptibility to type 2 diabetes mellitus (PSD) is associated with worse glycemic control and higher risk of cardiovascular events in stroke survivors.

**Design, Setting, and Participants:** We conducted a 3-stage genetic association study. First, we used a cross-sectional design and data from the UK Biobank to evaluate the relationship between PSD and glycemic control (enrollment took place between 2006 and 2010). Second, we used a cross-sectional design and data from the All of Us Research Program to replicate associations identified in the prior stage (enrollment between 2018 and 2022). Third, we used a prospective design and data from the Vitamin Intervention for Stroke Prevention (VISP) clinical trial to evaluate the relationship between PSD and post-stroke acute cardiovascular events (enrollment between 1997 and 2001). The present analyses were performed between May 2022 and August 2023.

**Exposures:** Low, intermediate, and high PSD modeled through categories of a polygenic risk score that included up to 462 independent DNA sequence variants associated with type 2 diabetes mellitus at genome-wide levels (p<5}10^-8^).

**Main Outcomes and Measures:** Hemoglobin A1c levels, treatment-resistant diabetes mellitus (defined as hemoglobin A1c ≥ 7.0% despite antidiabetic treatment), and risk of post-stroke acute cardiovascular events (stroke, myocardial infarction, or cardiovascular death).

**Results:** Stage 1 included 5,670 stroke survivors (mean age 61, 41% females), including 1,215 (21%) with known diabetes. Compared to stroke survivors with low PSD, those with high PSD had 35% higher hemoglobin A1c (beta 0.35, standard error [SE] 0.034; p <0.001) and 3 times the risk of having treatment-resistant diabetes (OR 3.40, 95%CI 2.52-6.24; p <0.001). These results pertaining to hemoglobin A1c remained significant when evaluating diabetics and non-diabetics separately (both tests p<0.05). Stage 2 replicated these results in 2,012 stroke survivors, including 447 (22.2%) with diabetes (mean age 64, 52% females, p <0.05 for all tests). Stage 3 included 1,750 stroke survivors (mean age 68, 34.8% females), including 441 (25.2%) with diabetes. Compared to stroke survivors with low PSD, those with high PSD had 50% higher risk of post-stroke cardiovascular events (hazard ratio 1.53, 95%CI 1.03, 2.28, p < 0.05).

**Conclusions and Relevance:** Among stroke survivors enrolled in 3 different studies, a higher PSD was associated with poorer glycemic control and higher risk of treatment-resistant diabetes and post-stroke vascular events. Given that millions of Americans are receiving diabetes-related polygenic risk data from direct-to-consumer companies, further research is needed to determine whether precision medicine strategies based on this information can improve the clinical management of these patients.

**KEY POINTS:** 

**Question:** Does polygenic susceptibility to type 2 diabetes mellitus lead to worse glycemic control and higher risk of cardiovascular events in stroke survivors?

**Findings:** Higher polygenic susceptibility to type 2 diabetes mellitus, modeled via polygenic risk scoring, was associated with higher levels of Hemoglobin A1c, higher risk of treatment resistant diabetes mellitus, and higher risk of recurrent cardiovascular events in stroke survivors.

**Meaning:** Polygenic susceptibility to type 2 diabetes is associated with worse clinical trajectories in stroke survivors. Given that millions of Americans are receiving diabetes-related polygenic risk data from direct-to-consumer companies, further research is needed to understand how this information could help post-stroke clinical management.

## INTRODUCTION

Stroke is the second leading cause of death and third leading cause of death and disability combined worldwide.^1^ Stroke survivors are a highly vulnerable group due to their increased risk of secondary diseases such as stroke recurrence, acute coronary events, cognitive decline, and dementia.^2–4^ One of the main determinants for post-stroke morbidity is diabetes mellitus, which is associated with a 2-fold increase in stroke recurrence and functional dependency.^5,6^ Even though the benefits of glycemic control on the morbidity and mortality of stroke survivors are well described, existing evidence indicates that up to 40% of stroke survivors have insufficiently controlled diabetes.^7^ The reasons why so many stroke survivors have inappropriate glycemic control are incompletely understood, although it is clear that multiple factors contribute to this scenario.^7^

Type 2 diabetes mellitus (T2DM) is a highly genetically determined disease with an estimated heritability (the proportion of a trait attributable to genetic variation) of 40% to 70%.^8,9^ In the setting of high-throughput genotyping technologies, this substantial genetic contribution has translated into numerous common genetic risk variants for T2DM.^10^ While each of these variants considered in isolation exerts a small effect in the risk of developing this disease, their aggregate burden, called polygenic susceptibility to diabetes (PSD), can lead to substantial increases in a person’s life-time risk of developing T2DM and associated vascular complications.^11,12^ Importantly, prior research showed that an elevated PSD has important clinical implications, leading to higher risk of first-ever acute vascular events.^12^

Despite the prominent role of genetic variation in T2DM, the clinical role of PSD in stroke survivors has not been studied. We therefore conducted a multistage genetic study that uses different study designs and data from several landmark studies to investigate whether PSD modifies the clinical trajectory of stroke survivors. We hypothesize that stroke survivors with an elevated PSD will have higher risk of uncontrolled diabetes as well as higher risk of post-stroke vascular events. These questions are important given that direct-to-consumer companies have returned polygenic risk information on several diseases, including T2DM, to millions of Americans and the National Institutes of Health have launched the All of Us Research Program to, among several goals, study the role of genomic information in precision medicine.^13^

## METHODS

### Study Design

We conducted a 3-stage genetic association study. First, we used a cross sectional design and data from the UK Biobank to evaluate the relationship between PSD and glycemic control (enrollment took place between 2006 and 2010).^14^ Second, we used a cross-sectional design and data from the All of Us Research Program to replicate associations identified in the prior stage (enrollment between 2018 and 2022).^13^ Third, we used a prospective design and data from the Vitamin Intervention for Stroke Prevention (VISP) clinical trial to evaluate the relationship between PSD and post-stroke acute cardiovascular events (enrollment took place between 1997 and 2001).^15–17^ Study protocols from these studies have been published previously, hence we provide brief summaries. Briefly, the UK Biobank is a large population study that has enrolled, and currently actively follows, ∼500,000 Britons.^14^ All of Us is an ongoing population study in the US that aims to enroll 1 million Americans with the ultimate goal of reducing health disparities and advancing precision medicine.^13^ VISP was a multicenter, double-blind, randomized, controlled clinical trial that randomized survivors of non-disabling stroke aged >35 with homocysteine levels above the 25th percentile to receive either low or high dose of daily multivitamin containing pyridoxine (Vitamin B_6_), folic acid (Vitamin B_9_), and cobalamin (Vitamin B_12_).^15–17^ All three studies have appropriate IRB oversight and participants provided written informed consent at enrollment.^13–17^ The present analyses were performed between May 2022 and August 2023.

### Qualifying event

The qualifying event for all analyses was prior ischemic or hemorrhagic stroke. From the UK Biobank, we included participants of European ancestry with a history of stroke at the time of enrollment. Qualifying events were ascertained using the UK Biobank’s algorithmically defined outcomes, which consists of self-reported data from the baseline study visit and electronic health record information from hospital admissions using previously validated International Classification of Diseases (ICD) codes (see Supplementary Table 1 for a list of the ICD codes used).^18^ In All of Us, we included participants of all ancestries with an ischemic or hemorrhagic stroke recorded in the baseline survey and health record data. VISP enrolled patients with a recent ischemic stroke and required both neuroimaging confirmation and evaluation by a board-certified neurologist to arrive at this diagnosis, thus we included all participants of European ancestry.^15–17^

### Genetic data

Detailed descriptions of DNA acquisition, genotyping procedures, quality control filters, and imputation pipelines for all 3 studies are available elsewhere.^14,19,20^ The genotyping chips used were the UK Biobank Axiom Array, Illumina Global Diversity Array, and the Illumina HumanOmni 1 for the UK Biobank, All of Us, and VISP, respectively. All data were quality controlled according to standard procedures in the field, including evaluation for batch effects, plate effects, departures from Hardy-Weinberg equilibrium, sex effects, array effects, case/control discordance, relatedness, and heterozygosity.^21^ The UK Biobank and All of Us used principal component analysis to account for population stratification and assign ancestry, while multidimensional scaling was utilized for VISP.^22,23^ Unobserved genotypes were imputed for all 3 studies using appropriate reference panels.^14,19,20^

### Exposure of interest

Our exposure of interest was PSD modeled through a polygenic risk score (PRS).^11^ For a given study participant, the PRS is the sum of the product of the risk allele counts for each variant multiplied by the allele’s reported effect on T2DM. To model PSD, we built a PRS using DNA sequence variants that involved a single nucleotide, had a minor allele frequency >1%, were independent of each other (r2 <0.1), were biallelic (involve two alleles only), and were associated with increased risk of T2DM at genome-wide levels (p<5}10^-8^).^10^ Using these criteria, the final PRS included 462 genetic risk variants in the UK Biobank and All of Us and 453 in VISP (Supplementary Table 2). These genetic variants were aligned to the GRCh37 assembly of the human genome and the effect allele was chosen to model increased risk of T2DM.^24^ PSD was then categorized as low (PRS <21st percentile), intermediate (PRS between the 21st and 80th percentiles), and high (>80th percentile).

### Outcome of interest

The outcomes of interest in Stages 1 (UK Biobank) and 2 (All of Us) were hemoglobin (Hb) A1c levels and treatment-resistant diabetes mellitus (defined as HbA1c ≥ 7.0% despite antidiabetic medication). HbA1c was measured during the baseline study visit (UK Biobank) or through electronic health record abstraction (All of Us). In the UK Biobank, antidiabetic medication was ascertained via dedicated research questionnaires and included metformin, rosiglitazone, glipizide, chlorpropamide, tolbutamide, acarbose, pioglitazone, rosiglitazone and insulin. Due to differences in medication assessment between the UK Biobank and All of Us, we could not replicate the analysis regarding treatment resistant diabetes in Stage 2. The outcome of interest in Stage 3 (VISP study) was a composite of recurrent stroke, incident coronary artery disease – defined as incident coronary artery bypass grafting or myocardial infarction – and death due to stroke or coronary cause. In VISP, these outcomes were assessed every three months for up to a two-year period, alternating between telephone contacts and in-clinic visits.

### Covariates

For the UK Biobank and All of Us participants, age in years was evaluated at enrollment. Sex and ancestry were determined using genetic information. Vascular risk factors and comorbidities were identified by combining baseline questionnaires, ICD 9/10 codes, and electronic health record data, as described for the ascertainment of qualifying events. For the VISP participants, age and vascular risk factors were obtained from the pre-filtered phenotypic data while sex and ancestry were determined using genetic information.

### Statistical analysis

We present discrete variables as counts (percentage [%]) and continuous variables as mean (SD) or median (interquartile range), as appropriate. All variables were evaluated to identify missing values and data entry errors. Unadjusted comparisons were made using chi-square tests for discrete variables and t or analysis of variance tests, as appropriate, for continuous variables. In stages 1 and 2, we used multivariable linear regression to evaluate changes in mean HbA1c levels as a function of PSD (low, intermediate, or high) while adjusting for age, sex, history of myocardial infarction, hypertension, hyperlipidemia, atrial fibrillation and smoking. In stratified analyses, we repeat<u>ed</u> these analyses in diabetics and non-diabetics. Similarly, in stage 1, we used multivariable logistic regression, adjusting for the same covariates, to evaluate changes in the mean odds of treatment-resistant diabetes as a function of PSD (low, intermediate or high). In stage 3, we used Cox proportional hazards models to evaluate the composite risk of vascular events as a function of PSD, adjusting for age, sex, hypertension, obesity, smoking, previous myocardial infarction, and VISP study treatment group. The proportional hazards assumption was tested by plotting the Schoenfeld residuals as a function of time. We declared statistical significance at p<0.05. We used PLINK to conduct quality control procedures,^23^ PRSice for polygenic risk scoring,^25^ and R (version 3.6)^26^ for all other analyses.

### Data availability statement

Data from the UK Biobank and All of Us are publicly available. UK Biobank Data was accessed using project application number 58743. Data from All of Us is publicly available at www.allofus.nih.gov and we used release version number 6, which includes data from all patients enrolled between May 30, 2017 and June 6, 2022. The phenotypic and genotypic data of VISP were acquired through dbGAP accession phs000343.v3.p1 and are available upon reasonable request from any qualified researcher.

## RESULTS

### Stage 1 – Discovery stage evaluating HbA1c and treatment-resistant diabetes mellitus

Of the total 488,203 participants with available genetic data enrolled in the UK Biobank, 6,997 had prevalent stroke at enrollment and were included in this study. Among these, 5,958 were of genetically-confirmed European ancestry. Of these, 287 (4.8%) were excluded due to missing HbA1c data and an additional 1 participant was excluded as an extreme outlier indicating either data entry or measurement error (Figure 1). The resulting analytical sample of Stage 1 consists of 5,670 stroke survivors (mean age 61 [SD 7] years; 2,315 [42%] were females). Of these, 1,397 (25%) were ischemic strokes, 982 (17%) were hemorrhagic strokes, and 3,291 (58%) were strokes of unknown mechanism. The mean time from stroke event to recruitment for this cohort was 7.85 years (SD 8.2). Table 1 gives an overview of the baseline characteristics.

**Figure 1:**
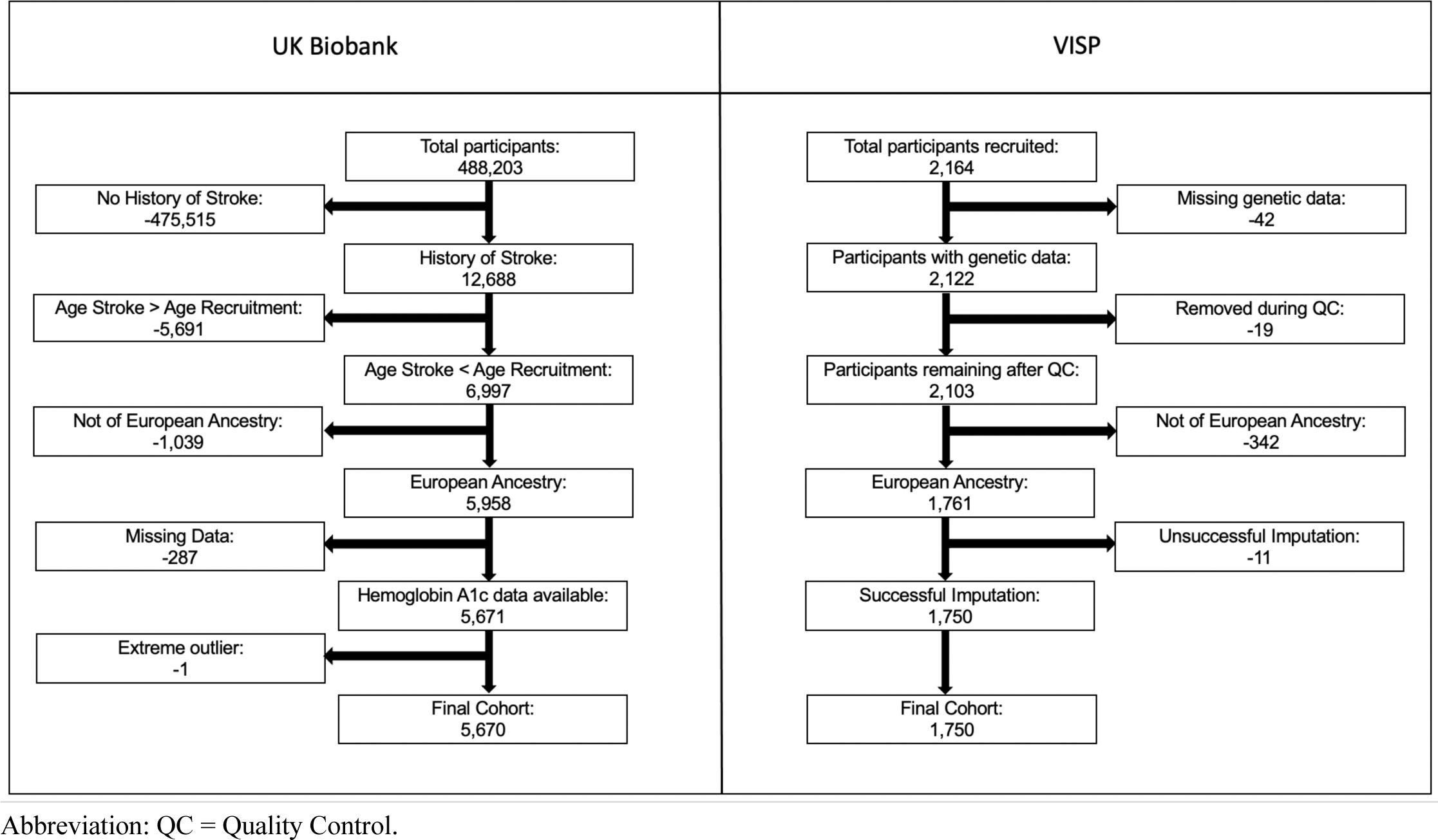
**Flowchart for cohort creation.**

**Table 1:**
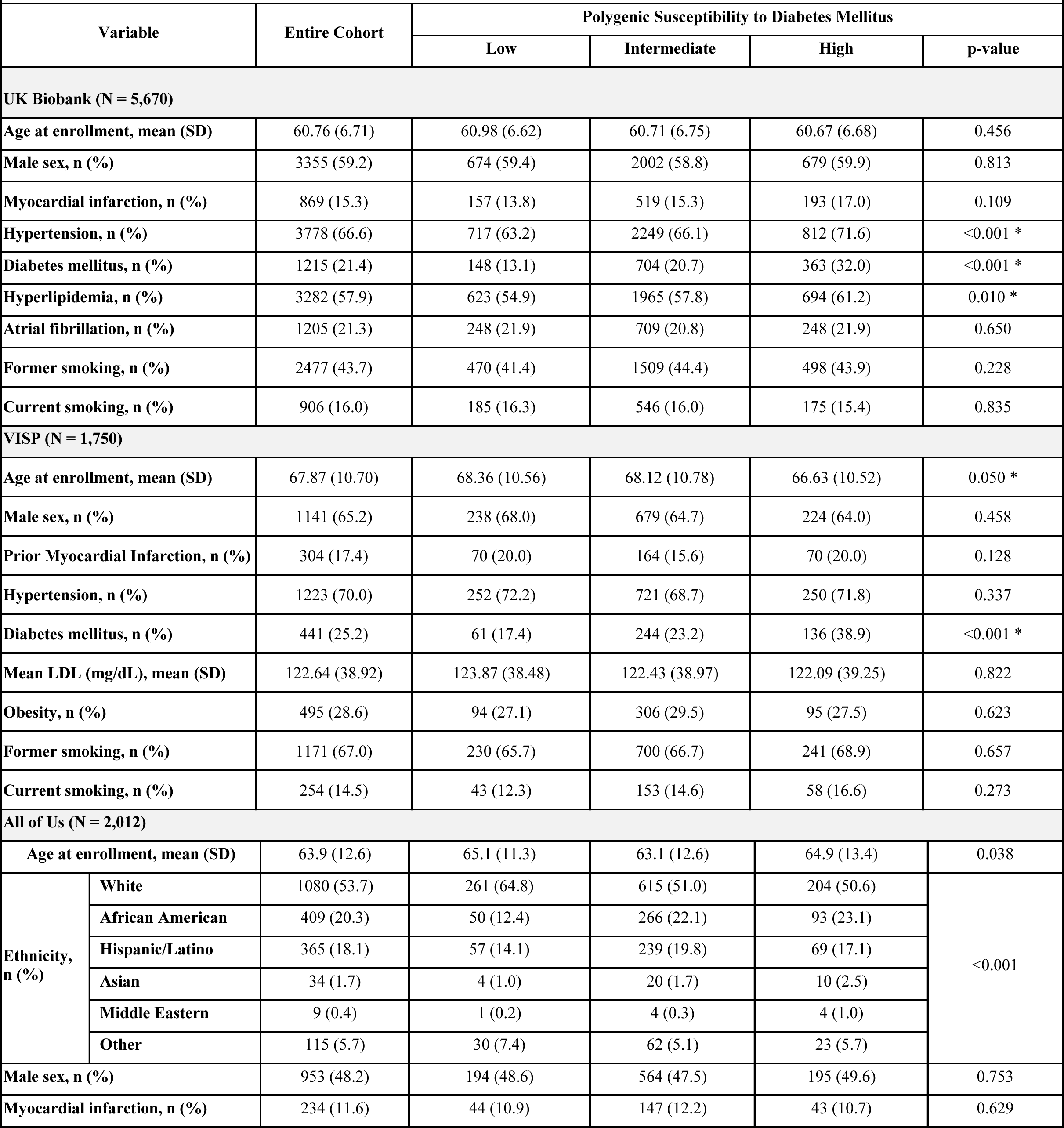
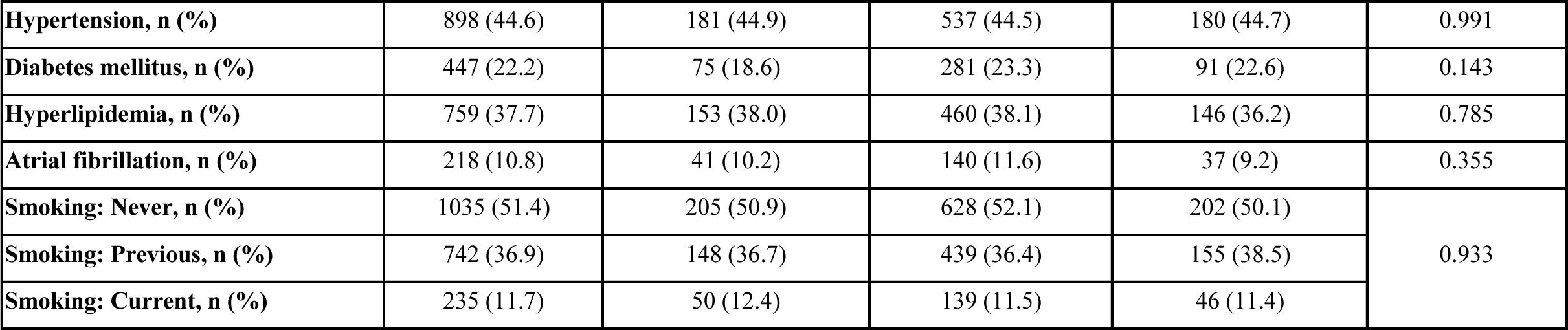
Baseline characteristics of study participants within each cohort.

|  |  |  |  |  |  |  |
| --- | --- | --- | --- | --- | --- | --- |
| Variable |  | Entire Cohort | Polygenic Susceptibility to Diabetes Mellitus |  |  |  |
|  |  |  | Low | Intermediate | High | p-value |
| UK Biobank (N = 5,670) |  |  |  |  |  |  |
| Age at enrollment, mean (SD) |  | 60.76 (6.71) | 60.98 (6.62) | 60.71 (6.75) | 60.67 (6.68) | 0.456 |
| Male sex, n (%) |  | 3355 (59.2) | 674 (59.4) | 2002 (58.8) | 679 (59.9) | 0.813 |
| Myocardial infarction, n (%) |  | 869 (15.3) | 157 (13.8) | 519 (15.3) | 193 (17.0) | 0.109 |
| Hypertension, n (%) |  | 3778 (66.6) | 717 (63.2) | 2249 (66.1) | 812 (71.6) | <0.001 * |
| Diabetes mellitus, n (%) |  | 1215 (21.4) | 148 (13.1) | 704 (20.7) | 363 (32.0) | <0.001 * |
| Hyperlipidemia, n (%) |  | 3282 (57.9) | 623 (54.9) | 1965 (57.8) | 694 (61.2) | 0.010 * |
| Atrial fibrillation, n (%) |  | 1205 (21.3) | 248 (21.9) | 709 (20.8) | 248 (21.9) | 0.650 |
| Former smoking, n (%) |  | 2477 (43.7) | 470 (41.4) | 1509 (44.4) | 498 (43.9) | 0.228 |
| Current smoking, n (%) |  | 906 (16.0) | 185 (16.3) | 546 (16.0) | 175 (15.4) | 0.835 |
| VISP (N = 1,750) |  |  |  |  |  |  |
| Age at enrollment, mean (SD) |  | 67.87 (10.70) | 68.36 (10.56) | 68.12 (10.78) | 66.63 (10.52) | 0.050 * |
| Male sex, n (%) |  | 1141 (65.2) | 238 (68.0) | 679 (64.7) | 224 (64.0) | 0.458 |
| Prior Myocardial Infarction, n (%) |  | 304 (17.4) | 70 (20.0) | 164 (15.6) | 70 (20.0) | 0.128 |
| Hypertension, n (%) |  | 1223 (70.0) | 252 (72.2) | 721 (68.7) | 250 (71.8) | 0.337 |
| Diabetes mellitus, n (%) |  | 441 (25.2) | 61 (17.4) | 244 (23.2) | 136 (38.9) | <0.001 * |
| Mean LDL (mg/dL), mean (SD) |  | 122.64 (38.92) | 123.87 (38.48) | 122.43 (38.97) | 122.09 (39.25) | 0.822 |
| Obesity, n (%) |  | 495 (28.6) | 94 (27.1) | 306 (29.5) | 95 (27.5) | 0.623 |
| Former smoking, n (%) |  | 1171 (67.0) | 230 (65.7) | 700 (66.7) | 241 (68.9) | 0.657 |
| Current smoking, n (%) |  | 254 (14.5) | 43 (12.3) | 153 (14.6) | 58 (16.6) | 0.273 |
| All of Us (N = 2,012) |  |  |  |  |  |  |
| Age at enrollment, mean (SD) |  | 63.9 (12.6) | 65.1 (11.3) | 63.1 (12.6) | 64.9 (13.4) | 0.038 |
| Ethnicity, n (%) | White | 1080 (53.7) | 261 (64.8) | 615 (51.0) | 204 (50.6) | <0.001 |
|  | African American | 409 (20.3) | 50 (12.4) | 266 (22.1) | 93 (23.1) |  |
|  | Hispanic/Latino | 365 (18.1) | 57 (14.1) | 239 (19.8) | 69 (17.1) |  |
|  | Asian | 34 (1.7) | 4 (1.0) | 20 (1.7) | 10 (2.5) |  |
|  | Middle Eastern | 9 (0.4) | 1 (0.2) | 4 (0.3) | 4 (1.0) |  |
|  | Other | 115 (5.7) | 30 (7.4) | 62 (5.1) | 23 (5.7) |  |
| Male sex, n (%) |  | 953 (48.2) | 194 (48.6) | 564 (47.5) | 195 (49.6) | 0.753 |
| Myocardial infarction, n (%) |  | 234 (11.6) | 44 (10.9) | 147 (12.2) | 43 (10.7) | 0.629 |

|  |  |  |  |  |  |
| --- | --- | --- | --- | --- | --- |
| **Hypertension, n (%)** | 898 (44.6) | 181 (44.9) | 537 (44.5) | 180 (44.7) | 0.991 |
| **Diabetes mellitus, n (%)** | 447 (22.2) | 75 (18.6) | 281 (23.3) | 91 (22.6) | 0.143 |
| **Hyperlipidemia, n (%)** | 759 (37.7) | 153 (38.0) | 460 (38.1) | 146 (36.2) | 0.785 |
| **Atrial fibrillation, n (%)** | 218 (10.8) | 41 (10.2) | 140 (11.6) | 37 (9.2) | 0.355 |
| **Smoking: Never, n (%)** | 1035 (51.4) | 205 (50.9) | 628 (52.1) | 202 (50.1) | 0.933 |
| **Smoking: Previous, n (%)** | 742 (36.9) | 148 (36.7) | 439 (36.4) | 155 (38.5) |  |
| **Smoking: Current, n (%)** | 235 (11.7) | 50 (12.4) | 139 (11.5) | 46 (11.4) |  |

The mean HbA1c was 5.7% (SD 0.83) and the overall prevalence of treatment-resistant diabetes mellitus was 4.8%. When comparing study participants with low versus high PSD via unadjusted analysis, the mean HbA1c was 5.51% (SD 0.60) versus 5.90% (SD 1.03), respectively (p<0.001). Similar differences were observed in adjusted multivariable analyses for HbA1c in all participants (beta 0.35, SE 0.034, p trend <0.001), participants without a history of diabetes mellitus (n=4,455, beta 0.096, SE 0.018, p trend <0.001), and participants with a history of diabetes mellitus (n=1,215, beta 0.412, SE 0.114, p trend <0.001, see Table 2). A higher PSD was also significantly associated with an increased occurrence of treatment resistant diabetes mellitus in stroke survivors. When comparing study participants with low versus high PSD in the unadjusted analysis, the proportion with treatment resistant diabetes was 2.2% versus 8.9% (p <0.001), respectively. We found a similar relationship in the adjusted multivariable analyses comparing high to low genetic risk strata (OR 3.40, 95%CI 2.52-6.24; p for trend <0.001, Table 2].

**Table 2:** Regression results between polygenic risk to diabetes and several outcomes in stroke survivors.

|  |  |  |  |  |  |  |
| --- | --- | --- | --- | --- | --- | --- |
| Cohort | Subgroup | Outcome | Analytical strategy | Genetic risk | Beta (SE) or OR (95%CI) or HR (95%CI) | Test for trend p |
| UK Biobank<br>N=5,670 | All participants | HbA1c | Linear regression | Low<br>Intermediate<br>High | Reference<br>0.17 (0.027)<br>0.35 (0.034) | <0.001 |
|  | Participants without diabetes | HbA1c | Linear regression | Low<br>Intermediate<br>High | Reference<br>0.065 (0.015)<br>0.096 (0.018) | <0.001 |
|  | Participants with diabetes | HbA1c | Linear regression | Low<br>Intermediate<br>High | Reference<br>0.23 (0.093)<br>0.41 (0.114) | <0.001 |
|  | Participants with diabetes | Treatment resistant | Logistic regression | Low<br>Intermediate<br>High | Reference<br>1.88 (1.24, 2.97)<br>3.40 (2.52, 6.24) | <0.001 |
| All of Us<br>N=2,012 | All participants | Hemoglobin A1c | Linear regression | Low<br>Intermediate<br>High | Reference<br>0.23 (0.14)<br>0.34 (0.17) | 0.04 |
| VISP<br>N=1,750 | All participants | Recurrent stroke or CAD | Cox regression | Low<br>Intermediate<br>High | Reference<br>1.11 (0.78, 1.57)<br>1.53 (1.03, 2.28) | 0.032 |

**Abbreviations: SE = Standard Error. OR = Odds Ratio. CI = Confidence Interval. HR = Hazard Ratio. CAD = Coronary Artery Disease.**

### Stage 2 – Replication of associations with HbA1c

Out of 372,397 participants enrolled in All of Us, 165,072 had chip array genomic data. Of these, 3,749 had an ischemic and 543 a hemorrhagic stroke. Of these, 1,012 participants had HbA1c measurements after stroke and were included in this replication analysis. When stratifying by ancestries, this cohort includes 1,080 (53.7%) white, 409 (20.3%) black or African American, 365 (18.1%) Hispanic or Latin American, 34 (1.7%) Asian, and 9 (0.4%) Middle Eastern or North African participants, and 115 (5.7%) participants of other heritage. When comparing study participants with low versus high PSD via unadjusted analysis, the mean HbA1c levels was 6.3% (SD 1.5) and 6.6% (SD 1.6), respectively (p=0.01). The differences remained significant in multivariable linear regression analyses (beta 0.34, SE 0.17; p 0.04, Table 2).

### Stage 3 – Association with post-stroke vascular events

Of the 2,122 study participants enrolled in VISP with genetic data available, there were 1,750 who were of genetically confirmed European ancestry remaining after standard quality control and imputation procedures [mean age 68 years (SD 11); 609 (35%) were females; see Table 1 and Figure 1]. After a mean follow up of 2.2 years, stroke survivors with low versus high PSD had a 50% higher risk of developing a post-stroke acute vascular event, including recurrent stroke, incident coronary artery disease, and/or death due to stroke or coronary cause (HR 1.53, 95%CI 1.03, 2.28; p<0.001, Table 2].

## DISCUSSION

We conducted a multi-stage genetic association study to study how PSD, or the aggregate contribution of known genetic risk variants for T2DM, modifies glycemic control and clinical outcomes in stroke survivors. The main motivation for the present study is the counterintuitive combination of lack of clear knowledge of the role of polygenic data in clinical practice and the rapidly growing number of Americans who have access to their genomic data via direct-to-consumer genotyping companies. We found that PSD, modeled via polygenic risk scoring, is associated with worse glycemic control, as indicated by higher HbA1c levels and higher risk of treatment-resistant diabetes in stroke survivors with higher PSD. We also found that that these findings have important clinical consequences, as stroke survivors with high PSD also had higher risk of post-stroke vascular events.

Extensive previous work demonstrated the important role of common genetic variation in T2DM, including the estimation of a high heritability,^8,9^ the identification of numerous genetic risk variants,^10^ and an association between high polygenic risk to T2DM and higher risk of vascular events, including stroke.^12^ However, knowledge on the effect of PSD on the clinical trajectories of stroke survivors is limited. In this study, we reinforce this relationship by verifying the clinical impact on elevated HbA1c and cardiovascular outcome after stroke. We also lay crucial groundwork to explore genetic predisposition to treatment-resistant diabetes in stroke patients. Our results can help to identify patients who are at increased risk of treatment failure, which enables physicians to individually allocate resources as opposed to a standardized “one size fits all” approach. The feasibility of tailored prevention strategies is ever-increasing, largely due to the decreased costs and increasing acceptance of genotyping.

Possible future applications of the findings described above are illustrated by an ancillary genetic study of the *Evaluation of Cardiovascular Outcomes After an Acute Coronary Syndrome During Treatment With Alirocumab* (ODYSSEY OUTCOMES) study, a randomized clinical trial that evaluated the PCSK9 inhibitor alirocumab in ∼18,000 patients with a recent acute coronary syndrome and low LDL cholesterol.^27^ Genetic analyses found that study participants with high versus low polygenic susceptibility to cardiovascular disease, modeled through PRSs, achieved significantly lower risk of recurrent vascular events with the study intervention.^28^ With additional evidence focused on other qualifying events like stroke, physicians could be able to individually adapt medication and other prevention strategies to the genetic profile of stroke patients. Furthermore, educating high-risk patients earlier in their clinical trajectories could improve timing and selection of interventions and sustain health-promoting lifestyle modifications in patients.^29^

The main strengths of our study are the large sample size and the multi-stage design, the latter allowing for independent replication and evaluation of similar and related questions in different populations. Our study has limitations. First, stroke cases within the UK Biobank and All of Us relied on validated International Classification of Diseases 9/10 codes extracted from electronic health records, which might have introduced misclassification. However, the positive predictive value of the ICD codes is estimated by the UK Biobank team to be around 85-90%.^30^ Second, HbA1c was measured only once, providing no insight on the long-term trajectories of diabetes control in these patients. Finally, even though part of the results could be replicated in the multi-ancestral population of All of Us, most of the studied stroke survivors were of European ancestry, limiting the portability of our findings to other race/ethnic groups.

In summary, PSD is associated with increased HbA1c levels, higher risk of treatment-resistant T2DM, higher risk of vascular events in stroke survivors. These findings support further research on the utility of genetically-based precision medicine strategies in populations of stroke survivors. These efforts will be greatly facilitated by the increasing accessibility and decreasing cost of high throughput genomic data.

## Supporting information

Diagnostic Codes and SNPs

## Data Availability

All data produced in the present work are contained in the manuscript

