## Supplementary material for "Polygenic Susceptibility to Diabetes and Poor Glycemic Control in Stroke Survivors": Diagnostic Codes and SNPs

**Supplementary Material for: Diabetes-Related Polygenic Risk Is Associated with Poor Glycemic Control and Higher Risk of Acute Cardiovascular Events in Stroke Survivors**

**Tables**

**Supplementary Table 1: Diagnostic codes used for stroke identification in UK Biobank Cohort.**

| | **ICD-9** | **ICD-10** |
|---|---|---|
| **Codes** | 430, 431, 434.0, 434.1, 434.9, 436.X | I60, I60.0, I60.1, I60.2, I60.3, I60.4, I60.5, I60.6, I60.7, I60.8, I60.9, I61, I61.0, I61.1, I61.2, I61.3, I61.4, I61.5, I61.6, I61.8, I61.9, I63, I63.0, I63.1, I63.2, I63.3, I63.4, I63.5, I63.6, I63.8, I63.9, I63.X, I64, I64.X |

**Supplementary Table 2: SNPs used to generate Polygenic Risk Scores.**

*Note that Asterisk and grey shading denotes SNP used in UK Biobank and All of Us Cohorts but not in VISP Cohort.

| CHR | BP | SNP | A1 | A2 | BETA | SE |
|---|---|---|---|---|---|---|
| 1 | 39855177 | rs61779284 | G | A | -7.03E-02 | 0.0048 |
| 1 | 214159256 | rs340874 | C | T | 5.43E-02 | 0.0039 |
| 1 | 51209148 | rs79090772 | C | T | -7.28E-02 | 0.0066 |
| 1 | 117532790 | rs1127215 | C | T | 4.12E-02 | 0.0039 |
| 1 | 120499573 | rs2453051 | C | A | 6.09E-02 | 0.0061 |
| 1 | 229672955 | rs348330 | G | A | 4.97E-02 | 0.0054 |
| 1 | 6672729 | rs11583755 | C | A | 3.69E-02 | 0.0041 |
| 1 | 118169463 | rs2282456 | G | A | -3.51E-02 | 0.0042 |
| 1 | 177889025 | rs539515 | C | A | 3.58E-02 | 0.0046 |
| 1 | 20707153 | rs10916780 | G | A | -3.42E-02 | 0.0046 |
| 1 | 64107284 | rs2269247 | C | T | 3.56E-02 | 0.0049 |
| 1 | 151017991 | rs145904381 | C | T | -1.61E-01 | 0.0221 |
| 1 | 155269776 | rs3020781 | G | A | 2.94E-02 | 0.0043 |
| 1 | 201849926 | rs41304257 | G | A | -3.18E-02 | 0.0047 |
| 1 | 235542023 | rs10737818 | G | A | 3.52E-02 | 0.0052 |
| 1 | 11317932 | rs7554251 | C | T | 2.97E-02 | 0.0045 |
| 1 | 206621028 | rs61817176 | C | A | -2.63E-02 | 0.004 |
| 1 | 29060898 | rs3753693 | C | T | 2.55E-02 | 0.004 |
| 1 | 205044339 | rs11240351 | G | A | 2.56E-02 | 0.004 |

| | | | | | | |
|---|---|---|---|---|---|---|
| 1 | 67010654 | rs4655617 | C | A | 2.56E-02 | 0.004 |
| 1 | 204490470 | rs12041243 | G | A | -2.93E-02 | 0.0047 |
| 1 | 16050470 | rs12746673 | C | A | 3.08E-02 | 0.0049 |
| 1 | 26396065 | rs9438610 | G | A | -3.13E-02 | 0.0051 |
| 1 | 200197538 | rs12128213 | G | A | -2.56E-02 | 0.0042 |
| 1 | 96404462 | rs10159026 | C | T | 2.73E-02 | 0.0046 |
| 1 | 65989878 | rs10889560 | C | A | -3.76E-02 | 0.0063 |
| 1 | 183004334 | rs4129858 | G | A | 2.23E-02 | 0.0038 |
| 1 | 179248952 | rs2816177 | G | A | 2.24E-02 | 0.0039 |
| 1 | 112289983 | rs197374 | C | T | -2.24E-02 | 0.0039 |
| 1 | 92048779 | rs4658234 | G | T | -2.95E-02 | 0.0051 |
| 1 | 33196120 | rs59020573 | G | A | -5.03E-02 | 0.0088 |
| 1 | 36789546 | rs12116935 | G | A | 2.42E-02 | 0.0043 |
| 1 | 62579891 | rs12140153 | G | T | 4.53E-02 | 0.0083 |
| 2 | 227105921 | rs2943650 | C | T | -8.05E-02 | 0.0042 |
| 2 | 43611883 | rs76675804 | C | T | -1.21E-01 | 0.0074 |
| 2 | 165508389 | rs10184004 | C | T | 6.36E-02 | 0.0041 |
| 2 | 65279414 | rs2723065 | G | A | -4.22E-02 | 0.0039 |
| **2** | **653874** | ***rs10188334** | **C** | **T** | **5.44E-02** | **0.0053** |
| 2 | 121337196 | rs9308614 | G | A | -5.70E-02 | 0.0058 |
| 2 | 58975143 | rs12986742 | C | T | 3.53E-02 | 0.0038 |
| 2 | 181618654 | rs6741676 | G | A | -3.24E-02 | 0.0042 |
| 2 | 146350724 | rs6716394 | G | A | 2.75E-02 | 0.0037 |

| | | | | | | |
|---|---|---|---|---|---|---|
| 2 | 161333872 | rs6710938 | C | A | -3.42E-02 | 0.0046 |
| 2 | 111887754 | rs113135335 | G | T | -5.33E-02 | 0.0073 |
| 2 | 152198598 | rs3845843 | C | T | -2.64E-02 | 0.0038 |
| 2 | 25635771 | rs34845373 | G | A | -3.34E-02 | 0.0048 |
| 2 | 166610827 | rs13406280 | C | T | 2.69E-02 | 0.0039 |
| 2 | 196952010 | rs6712905 | C | T | 2.79E-02 | 0.0042 |
| 2 | 67622243 | rs4671799 | G | A | -2.62E-02 | 0.004 |
| 2 | 203235139 | rs6714523 | G | A | -4.01E-02 | 0.0061 |
| 2 | 228971884 | rs13415288 | C | T | 2.62E-02 | 0.004 |
| 2 | 208870017 | rs34329895 | G | A | -2.55E-02 | 0.004 |
| 2 | 213829721 | rs4673712 | C | T | -2.69E-02 | 0.0045 |
| 2 | 16238001 | rs28758542 | G | A | -2.56E-02 | 0.0042 |
| 2 | 55157914 | rs5010712 | G | A | 3.00E-02 | 0.0051 |
| 2 | 112823114 | rs74677818 | C | T | -3.57E-02 | 0.006 |
| 2 | 37204168 | rs77424687 | C | T | 2.74E-02 | 0.0046 |
| 2 | 18707873 | rs11096542 | G | A | -2.54E-02 | 0.0043 |
| 2 | 26192802 | rs72803684 | C | T | -6.66E-02 | 0.0115 |
| 2 | 212274937 | rs3828242 | G | A | -2.52E-02 | 0.0043 |
| 2 | 100598726 | rs34506349 | G | A | 6.65E-02 | 0.0116 |
| 2 | 175197545 | rs12992995 | C | A | 2.72E-02 | 0.0048 |
| 2 | 179650954 | rs6715901 | G | A | 2.30E-02 | 0.004 |
| 2 | 219859171 | rs113414093 | G | A | -7.24E-02 | 0.0128 |
| 2 | 205375909 | rs4482463 | C | A | 3.16E-02 | 0.0056 |

| | | | | | | |
|---|---|---|---|---|---|---|
| 2 | 163623932 | rs305686 | C | T | 2.94E-02 | 0.0053 |
| 2 | 105165674 | rs10469860 | G | A | -2.38E-02 | 0.0043 |
| 2 | 86707504 | rs4832290 | C | T | -2.67E-02 | 0.0049 |
| 2 | 219168432 | rs1877712 | G | A | 2.11E-02 | 0.0039 |
| 3 | 185534482 | rs9859406 | G | A | -1.12E-01 | 0.004 |
| 3 | 123065778 | rs11708067 | G | A | -8.04E-02 | 0.005 |
| 3 | 12329783 | rs17036160 | C | T | 1.01E-01 | 0.0065 |
| 3 | 23457080 | rs13094957 | C | T | -6.37E-02 | 0.0047 |
| 3 | 187741842 | rs6777684 | G | A | 5.48E-02 | 0.0044 |
| 3 | 63948566 | rs13434089 | C | T | -6.07E-02 | 0.0052 |
| 3 | 170724883 | rs8192675 | C | T | -4.52E-02 | 0.0042 |
| 3 | 186665645 | rs3887925 | C | T | -4.01E-02 | 0.0038 |
| 3 | 124921457 | rs9873519 | C | T | -3.73E-02 | 0.0038 |
| 3 | 64703394 | rs66815886 | G | T | 4.00E-02 | 0.0042 |
| 3 | 152084243 | rs7633673 | G | A | 3.06E-02 | 0.0038 |
| 3 | 195831237 | rs9872347 | C | T | 3.18E-02 | 0.0042 |
| **3** | **160153305** | ***rs7629** | **G** | **A** | **-3.11E-02** | **0.0042** |
| 3 | 136069472 | rs667920 | G | T | -3.24E-02 | 0.0045 |
| 3 | 115063672 | rs7645613 | C | T | 3.40E-02 | 0.0048 |
| 3 | 138055136 | rs6766859 | C | T | 2.75E-02 | 0.004 |
| 3 | 9514016 | rs3872707 | G | A | -3.70E-02 | 0.0054 |
| 3 | 15706124 | rs924753 | G | A | 2.63E-02 | 0.0039 |
| 3 | 86756871 | rs6549112 | G | T | -2.68E-02 | 0.004 |

| | | | | | | |
|---|---|---|---|---|---|---|
| 3 | 173107443 | rs247975 | C | T | 2.54E-02 | 0.0038 |
| 3 | 183738460 | rs2872246 | C | A | -2.66E-02 | 0.004 |
| 3 | 72803590 | rs9814945 | C | T | -3.16E-02 | 0.0048 |
| 3 | 129293256 | rs2255703 | C | T | -2.56E-02 | 0.0039 |
| 3 | 184882015 | rs61579137 | G | A | -3.09E-02 | 0.0047 |
| 3 | 149221563 | rs28712435 | C | T | 2.54E-02 | 0.0039 |
| 3 | 168226052 | rs13099581 | C | T | 3.71E-02 | 0.0058 |
| 3 | 47693664 | rs62262091 | C | T | -5.38E-02 | 0.0084 |
| 3 | 131750844 | rs9857204 | G | A | -2.72E-02 | 0.0043 |
| 3 | 35670150 | rs1470560 | G | A | -2.45E-02 | 0.004 |
| 3 | 121961461 | rs13059382 | G | T | 3.56E-02 | 0.0059 |
| 3 | 54828827 | rs76263492 | G | T | -6.75E-02 | 0.0114 |
| 3 | 93981060 | rs978444 | G | T | 2.29E-02 | 0.0039 |
| 3 | 77671721 | rs2272163 | C | A | 2.35E-02 | 0.004 |
| 3 | 28731810 | rs9869477 | G | A | 2.26E-02 | 0.0038 |
| 3 | 88130136 | rs73146095 | C | T | 4.06E-02 | 0.007 |
| 3 | 113288430 | rs11929640 | G | A | -2.45E-02 | 0.0042 |
| 3 | 173710695 | rs59489841 | C | T | 2.35E-02 | 0.0042 |
| 4 | 1240299 | rs730831 | G | T | -8.78E-02 | 0.0072 |
| 4 | 153520475 | rs6813195 | C | T | 4.63E-02 | 0.004 |
| 4 | 45186139 | rs10938398 | G | A | -4.45E-02 | 0.0039 |
| 4 | 1784403 | rs56337234 | C | T | 4.26E-02 | 0.0041 |
| 4 | 720681 | rs73221116 | G | A | -9.55E-02 | 0.0106 |

| | | | | | | |
|---|---|---|---|---|---|---|
| 4 | 185716100 | rs55691245 | G | A | 5.61E-02 | 0.0064 |
| 4 | 71835822 | rs7674402 | G | A | -5.23E-02 | 0.0064 |
| 4 | 18044357 | rs2169033 | C | T | -3.25E-02 | 0.0041 |
| 4 | 157652753 | rs28819812 | C | A | 3.30E-02 | 0.0042 |
| 4 | 103895317 | rs7659468 | G | T | -2.87E-02 | 0.0037 |
| 4 | 106048291 | rs17035289 | C | T | 3.57E-02 | 0.0048 |
| 4 | 52798624 | rs1996617 | C | T | 2.89E-02 | 0.004 |
| 4 | 140906390 | rs12505942 | C | T | -2.94E-02 | 0.0042 |
| 4 | 83584496 | rs993380 | G | A | -2.72E-02 | 0.0039 |
| 4 | 49067323 | rs2605281 | G | A | -2.83E-02 | 0.0041 |
| 4 | 96114385 | rs3755879 | G | A | -2.73E-02 | 0.0042 |
| 4 | 76496817 | rs6835992 | G | A | 2.87E-02 | 0.0044 |
| 4 | 186580062 | rs35901985 | G | A | -3.32E-02 | 0.0053 |
| 4 | 95114572 | rs28408270 | G | T | 2.39E-02 | 0.0039 |
| 4 | 121765788 | rs4833687 | C | T | -2.67E-02 | 0.0045 |
| 4 | 156697784 | rs2125799 | C | T | 2.44E-02 | 0.0041 |
| 4 | 129024273 | rs4834232 | C | T | -2.72E-02 | 0.0047 |
| 4 | 20265535 | rs7664347 | C | T | -2.34E-02 | 0.0041 |
| 4 | 77017680 | rs4440243 | C | T | -2.46E-02 | 0.0043 |
| 4 | 130786346 | rs2952858 | G | A | -2.37E-02 | 0.0041 |
| 4 | 77533939 | rs56281442 | G | A | -2.50E-02 | 0.0044 |
| 4 | 44503503 | rs34617913 | G | A | -2.39E-02 | 0.0043 |
| 4 | 3241845 | rs362307 | C | T | -4.80E-02 | 0.0087 |

| | | | | | | |
|---|---|---|---|---|---|---|
| 5 | 55807370 | rs464605 | C | T | -7.00E-02 | 0.0041 |
| 5 | 102586407 | rs75432112 | G | A | -1.32E-01 | 0.0104 |
| 5 | 76435004 | rs7732130 | G | A | 5.54E-02 | 0.0047 |
| 5 | 53272664 | rs4865796 | G | A | -4.81E-02 | 0.0042 |
| 5 | 158029734 | rs1650505 | G | A | -4.09E-02 | 0.0045 |
| 5 | 78546293 | rs2591392 | G | A | 3.06E-02 | 0.0039 |
| 5 | 133861663 | rs329118 | C | T | -2.81E-02 | 0.0038 |
| 5 | 51763665 | rs12187734 | C | T | 2.87E-02 | 0.0039 |
| 5 | 122650885 | rs144052331 | C | T | -7.09E-02 | 0.0098 |
| 5 | 50145266 | rs152839 | C | T | 2.81E-02 | 0.0039 |
| 5 | 44644006 | rs4479849 | G | A | -2.58E-02 | 0.0039 |
| 5 | 176679407 | rs4343858 | G | A | 2.95E-02 | 0.0045 |
| 5 | 67714246 | rs4976033 | G | A | 2.51E-02 | 0.0039 |
| 5 | 95850250 | rs261967 | C | A | 2.35E-02 | 0.0038 |
| 5 | 87697533 | rs6870983 | C | T | 2.87E-02 | 0.0049 |
| 5 | 58132702 | rs2662390 | C | T | -2.76E-02 | 0.0048 |
| 5 | 87102981 | rs73167517 | G | T | -4.11E-02 | 0.0072 |
| 5 | 137431501 | rs217256 | C | T | 2.10E-02 | 0.0037 |
| 5 | 151324600 | rs302395 | G | T | 2.32E-02 | 0.0041 |
| 5 | 46204748 | rs8188241 | G | A | 2.32E-02 | 0.0041 |
| 5 | 170683134 | rs2913873 | G | A | -3.16E-02 | 0.0056 |
| 5 | 54986775 | rs6897117 | C | T | -2.34E-02 | 0.0042 |
| 5 | 36084426 | rs114136102 | C | T | 6.31E-02 | 0.0114 |

| | | | | | | |
|---|---|---|---|---|---|---|
| 5 | 180226516 | rs6885157 | G | T | 3.72E-02 | 0.0068 |
| 6 | 137295352 | rs2876354 | C | T | 4.65E-02 | 0.0038 |
| 6 | 31134888 | rs3130931 | C | T | 4.82E-02 | 0.0041 |
| 6 | 7231843 | rs9379084 | G | A | 7.15E-02 | 0.0062 |
| 6 | 32138545 | rs3130283 | C | A | 6.17E-02 | 0.0057 |
| 6 | 126792095 | rs11759026 | G | A | 5.85E-02 | 0.0057 |
| 6 | 50788778 | rs3798519 | C | A | 4.74E-02 | 0.0047 |
| 6 | 127412728 | rs12192275 | G | A | 4.82E-02 | 0.005 |
| 6 | 164133001 | rs4709746 | C | T | 5.65E-02 | 0.0059 |
| 6 | 117996631 | rs80196932 | C | T | -4.79E-02 | 0.0053 |
| 6 | 160770918 | rs501470 | G | T | -3.20E-02 | 0.0038 |
| 6 | 153440770 | rs7758002 | G | T | -3.17E-02 | 0.0039 |
| 6 | 39282371 | rs34247110 | G | A | -3.05E-02 | 0.0038 |
| 6 | 143056556 | rs9390022 | C | T | -3.06E-02 | 0.0041 |
| 6 | 33552707 | rs75080135 | C | A | -3.57E-02 | 0.0049 |
| 6 | 107445266 | rs7752666 | C | T | 3.05E-02 | 0.0043 |
| 6 | 131926334 | rs2608953 | C | T | -3.48E-02 | 0.005 |
| 6 | 138864489 | rs7742292 | C | T | 2.61E-02 | 0.004 |
| 6 | 53789830 | rs9370243 | G | T | -3.94E-02 | 0.0061 |
| 6 | 40409243 | rs34298980 | C | T | -3.54E-02 | 0.0055 |
| 6 | 51413013 | rs16881572 | G | A | -6.81E-02 | 0.011 |
| 6 | 111738793 | rs55812705 | C | T | -3.01E-02 | 0.005 |
| **6** | **35259397** | ***rs33959228** | **C** | **T** | **9.15E-02** | **0.0155** |

| | | | | | | |
|---|---|---|---|---|---|---|
| 6 | 126061502 | rs7758115 | G | A | -2.32E-02 | 0.0039 |
| 6 | 34236973 | rs2780215 | G | A | -5.31E-02 | 0.0091 |
| 6 | 19751516 | rs10806906 | C | T | 2.23E-02 | 0.0039 |
| 6 | 130265266 | rs35164294 | G | T | -5.25E-02 | 0.0092 |
| 6 | 29816421 | rs2394186 | G | A | -2.71E-02 | 0.0048 |
| 6 | 26211146 | rs9358912 | G | T | 2.49E-02 | 0.0045 |
| 6 | 64163807 | rs9449295 | C | T | 2.16E-02 | 0.0039 |
| 6 | 36911274 | rs72846863 | G | A | 3.35E-02 | 0.0061 |
| 7 | 28194397 | rs860262 | C | A | 7.47E-02 | 0.0039 |
| 7 | 156948648 | rs6946660 | C | T | -4.72E-02 | 0.0039 |
| 7 | 44223721 | rs730497 | G | A | -5.52E-02 | 0.005 |
| 7 | 69649683 | rs6975279 | C | A | -4.31E-02 | 0.0042 |
| 7 | 30728452 | rs917195 | C | T | 4.24E-02 | 0.0046 |
| 7 | 150537635 | rs62492368 | G | A | -3.11E-02 | 0.0041 |
| 7 | 12269593 | rs13237518 | C | A | -2.83E-02 | 0.0038 |
| 7 | 13886654 | rs7787720 | C | T | -2.85E-02 | 0.0039 |
| 7 | 18331915 | rs583769 | G | A | -3.07E-02 | 0.0043 |
| 7 | 140522073 | rs60251368 | G | A | 5.49E-02 | 0.0078 |
| 7 | 102976385 | rs187653072 | C | T | 9.67E-02 | 0.0138 |
| 7 | 23884697 | rs2188848 | G | A | -3.25E-02 | 0.0048 |
| 7 | 48839003 | rs12539264 | G | A | 2.90E-02 | 0.0045 |
| 7 | 100313420 | rs506597 | G | A | 4.10E-02 | 0.0064 |
| 7 | 50577968 | rs73121277 | C | T | 2.60E-02 | 0.0042 |

| | | | | | | |
|---|---|---|---|---|---|---|
| 7 | 15926228 | rs38221 | C | T | -2.86E-02 | 0.0047 |
| 7 | 4683572 | rs62452060 | G | A | -3.54E-02 | 0.0059 |
| 7 | 74076493 | rs13238568 | G | A | 2.48E-02 | 0.0041 |
| 7 | 55802063 | rs6972291 | C | T | 2.83E-02 | 0.0048 |
| 7 | 77047102 | rs12669521 | G | A | -2.63E-02 | 0.0044 |
| 7 | 40816653 | rs17439448 | C | T | -3.93E-02 | 0.0067 |
| 7 | 142607301 | rs4252505 | G | A | 4.13E-02 | 0.007 |
| 7 | 36742886 | rs6978327 | C | T | -2.33E-02 | 0.004 |
| 7 | 45116468 | rs3735491 | C | A | -2.50E-02 | 0.0043 |
| **7** | **93118736** | ***rs10262104** | **C** | **T** | **2.20E-02** | **0.0039** |
| 7 | 104516274 | rs73184014 | G | A | -2.98E-02 | 0.0052 |
| 7 | 117495667 | rs6976111 | C | A | -2.58E-02 | 0.0046 |
| 7 | 147658539 | rs1922879 | G | A | 2.30E-02 | 0.0041 |
| 7 | 131574608 | rs12667919 | C | A | -2.56E-02 | 0.0047 |
| 7 | 148429806 | rs243513 | G | A | 2.32E-02 | 0.0043 |
| 7 | 149238823 | rs62490267 | C | T | 3.10E-02 | 0.0057 |
| 8 | 118184783 | rs13266634 | C | T | 1.02E-01 | 0.0041 |
| 8 | 41508577 | rs13262861 | C | A | 1.02E-01 | 0.0054 |
| 8 | 95967372 | rs10808671 | G | A | -3.58E-02 | 0.0038 |
| 8 | 145525277 | rs13268508 | C | T | -3.76E-02 | 0.0041 |
| 8 | 9974584 | rs60384372 | G | A | -3.52E-02 | 0.0042 |
| 8 | 11069960 | rs2409742 | C | T | 3.41E-02 | 0.0042 |
| 8 | 8721473 | rs4382480 | G | A | -3.06E-02 | 0.0042 |

| | | | | | | |
|---|---|---|---|---|---|---|
| 8 | 129568078 | rs1561927 | C | T | 3.32E-02 | 0.0046 |
| 8 | 36858483 | rs13365225 | G | A | 3.27E-02 | 0.0046 |
| 8 | 30852826 | rs2725370 | C | T | -3.19E-02 | 0.0046 |
| 8 | 57496064 | rs3887059 | G | A | -2.95E-02 | 0.0043 |
| 8 | 9265105 | rs17662402 | C | T | -6.70E-02 | 0.01 |
| 8 | 105662373 | rs112515915 | G | A | -5.62E-02 | 0.0085 |
| 8 | 12643055 | rs12056338 | G | T | -2.51E-02 | 0.004 |
| 8 | 22492103 | rs6558173 | G | T | -2.45E-02 | 0.004 |
| 8 | 19830921 | rs10096633 | C | T | 3.30E-02 | 0.0054 |
| 8 | 25871721 | rs11998023 | G | T | 3.24E-02 | 0.0053 |
| 8 | 14148990 | rs35753840 | C | A | 2.54E-02 | 0.0042 |
| 8 | 34502571 | rs4463416 | C | A | -2.37E-02 | 0.0041 |
| 8 | 97138738 | rs510062 | G | A | -2.20E-02 | 0.0039 |
| 8 | 28095939 | rs11994255 | C | T | -2.39E-02 | 0.0043 |
| 8 | 135775546 | rs4294149 | C | T | -2.17E-02 | 0.0039 |
| 8 | 37397803 | rs12680217 | C | T | -3.97E-02 | 0.0071 |
| 8 | 128711742 | rs17772814 | G | A | 6.85E-02 | 0.0124 |
| 8 | 74568099 | rs28792187 | G | A | 4.14E-02 | 0.0076 |
| 9 | 22134094 | rs10811661 | C | T | -1.47E-01 | 0.0048 |
| 9 | 139248082 | rs28642213 | G | A | 8.22E-02 | 0.0047 |
| 9 | 84308948 | rs2796441 | G | A | 6.09E-02 | 0.0039 |
| **9** | **136149500** | ***rs529565** | **C** | **T** | **4.15E-02** | **0.0039** |
| 9 | 28410683 | rs1412234 | C | T | 3.97E-02 | 0.0042 |

| | | | | | | |
|---|---|---|---|---|---|---|
| 9 | 81907986 | rs67269808 | G | A | -7.31E-02 | 0.0078 |
| 9 | 34074476 | rs12001437 | C | T | 3.01E-02 | 0.0038 |
| 9 | 96915002 | rs10993072 | C | T | -3.15E-02 | 0.0041 |
| 9 | 19074538 | rs12380322 | G | A | 2.95E-02 | 0.004 |
| 9 | 23358495 | rs7029718 | G | A | -2.73E-02 | 0.0039 |
| 9 | 125689694 | rs10818763 | C | T | 3.58E-02 | 0.0051 |
| 9 | 136890704 | rs379417 | G | A | -2.78E-02 | 0.0042 |
| 9 | 116943357 | rs1431819 | G | A | 2.94E-02 | 0.0046 |
| 9 | 119252277 | rs1885234 | G | T | 2.44E-02 | 0.0039 |
| 9 | 35749014 | rs1570247 | G | A | 2.44E-02 | 0.0039 |
| 9 | 20790622 | rs2150999 | C | T | 2.43E-02 | 0.0039 |
| 9 | 97795421 | rs6479591 | G | A | -3.26E-02 | 0.0054 |
| 9 | 111938268 | rs10119430 | G | A | 2.79E-02 | 0.0046 |
| 9 | 14141703 | rs73642097 | G | A | 2.91E-02 | 0.0049 |
| 9 | 133786652 | rs6597649 | C | T | -2.34E-02 | 0.0039 |
| 9 | 134868417 | rs9411425 | G | T | 2.36E-02 | 0.004 |
| 9 | 81344701 | rs1929883 | G | A | 2.75E-02 | 0.005 |
| 9 | 29089437 | rs13288108 | C | A | 2.74E-02 | 0.005 |
| 10 | 94462882 | rs1111875 | C | T | 9.28E-02 | 0.0038 |
| 10 | 12307894 | rs11257655 | C | T | -9.46E-02 | 0.0056 |
| 10 | 80947438 | rs697239 | C | T | -5.87E-02 | 0.0038 |
| 10 | 124193181 | rs2280141 | G | T | -4.55E-02 | 0.0048 |
| 10 | 71321279 | rs177045 | G | A | 3.35E-02 | 0.004 |

| | | | | | | |
|---|---|---|---|---|---|---|
| 10 | 99091369 | rs945187 | G | A | 3.10E-02 | 0.0039 |
| 10 | 70382179 | rs10998338 | G | A | -3.08E-02 | 0.0039 |
| 10 | 112678657 | rs7895872 | G | T | -3.06E-02 | 0.0041 |
| 10 | 89722731 | rs36062478 | C | T | 3.89E-02 | 0.0054 |
| 10 | 122930568 | rs7071036 | C | T | -5.10E-02 | 0.0073 |
| 10 | 64970928 | rs111765639 | G | A | 5.81E-02 | 0.0084 |
| 10 | 88117318 | rs11201992 | C | A | 2.51E-02 | 0.0038 |
| 10 | 101912194 | rs1408579 | C | T | 2.75E-02 | 0.0042 |
| 10 | 33997227 | rs71495046 | C | A | 3.99E-02 | 0.0063 |
| 10 | 100421841 | rs524903 | G | A | 4.28E-02 | 0.0067 |
| 10 | 44027356 | rs3122231 | C | T | 2.71E-02 | 0.0044 |
| 10 | 121660400 | rs11199116 | C | A | 3.06E-02 | 0.005 |
| 10 | 75598099 | rs2633311 | C | T | 2.37E-02 | 0.004 |
| 10 | 13540869 | rs11258422 | C | A | 2.41E-02 | 0.0041 |
| 10 | 125226178 | rs705145 | C | A | -2.34E-02 | 0.0039 |
| 10 | 63717113 | rs146716733 | C | T | 4.51E-02 | 0.0079 |
| 10 | 103065789 | rs620191 | G | T | -2.34E-02 | 0.0041 |
| 10 | 72648336 | rs827237 | C | T | -2.94E-02 | 0.0053 |
| 11 | 2858546 | rs2237897 | C | T | 2.20E-01 | 0.0068 |
| 11 | 2197286 | rs4929965 | G | A | -6.76E-02 | 0.0042 |
| 11 | 17418477 | rs757110 | C | A | 5.99E-02 | 0.0039 |
| 11 | 69463273 | rs3918298 | G | A | 1.04E-01 | 0.0101 |
| 11 | 128234144 | rs10750397 | G | A | -4.00E-02 | 0.0042 |

| | | | | | | |
|---|---|---|---|---|---|---|
| 11 | 65326154 | rs12789028 | G | A | -4.59E-02 | 0.005 |
| 11 | 8677063 | rs7941510 | C | A | 3.53E-02 | 0.0042 |
| **11** | **47857253** | ***rs3816605** | **C** | **T** | **-3.18E-02** | **0.0043** |
| 11 | 45858584 | rs12419690 | G | A | 2.98E-02 | 0.004 |
| **11** | **32956492** | ***rs62618693** | **C** | **T** | **8.19E-02** | **0.0111** |
| 11 | 43878459 | rs35251247 | G | A | -3.09E-02 | 0.0042 |
| 11 | 50110597 | rs11561066 | C | T | 4.51E-02 | 0.0065 |
| 11 | 58128015 | rs7483027 | C | T | -2.67E-02 | 0.0041 |
| 11 | 9856015 | rs76789970 | C | T | -3.87E-02 | 0.006 |
| 11 | 34908780 | rs2956092 | C | T | -2.54E-02 | 0.004 |
| 11 | 76230357 | rs2513505 | C | A | -2.40E-02 | 0.0038 |
| 11 | 117693255 | rs529623 | C | T | -2.24E-02 | 0.0038 |
| 11 | 55588216 | rs116861182 | C | A | 6.15E-02 | 0.0105 |
| 11 | 75464344 | rs11236524 | C | T | 3.63E-02 | 0.0064 |
| 11 | 118953202 | rs7127212 | C | T | -2.27E-02 | 0.0041 |
| 11 | 64100776 | rs1662185 | G | A | -2.26E-02 | 0.0041 |
| 11 | 20952237 | rs16907058 | G | A | -4.61E-02 | 0.0084 |
| 11 | 95710493 | rs7130522 | C | A | 2.22E-02 | 0.0041 |
| 12 | 4384844 | rs76895963 | G | T | -4.22E-01 | 0.0202 |
| 12 | 66216162 | rs2257883 | G | A | -7.10E-02 | 0.0049 |
| 12 | 108629780 | rs1426371 | G | A | 4.29E-02 | 0.0045 |
| 12 | 118406696 | rs79310463 | C | T | -5.00E-02 | 0.0053 |
| 12 | 26463174 | rs11048457 | G | A | 3.85E-02 | 0.0043 |

| | | | | | | |
|---|---|---|---|---|---|---|
| 12 | 133070294 | rs11614914 | C | T | -3.94E-02 | 0.0044 |
| 12 | 71520761 | rs10879261 | G | T | 3.41E-02 | 0.0038 |
| 12 | 124428331 | rs4930726 | C | T | -3.53E-02 | 0.0041 |
| 12 | 123736084 | rs10773000 | G | T | 3.40E-02 | 0.0041 |
| 12 | 57146069 | rs2277339 | G | T | 4.37E-02 | 0.0056 |
| 12 | 97849120 | rs6538805 | C | T | -3.03E-02 | 0.0039 |
| 12 | 50263148 | rs7132908 | G | A | -2.97E-02 | 0.004 |
| 12 | 41863393 | rs2730827 | C | T | -2.79E-02 | 0.0038 |
| 12 | 33410855 | rs10844519 | G | T | 3.00E-02 | 0.0042 |
| 12 | 43046449 | rs11181613 | C | A | 4.18E-02 | 0.006 |
| 12 | 133730500 | rs7970687 | C | T | -2.60E-02 | 0.0039 |
| 12 | 95928113 | rs11108094 | C | A | -5.71E-02 | 0.0088 |
| 12 | 12871099 | rs2066827 | G | T | 3.21E-02 | 0.0051 |
| 12 | 20591332 | rs7134150 | G | A | -4.43E-02 | 0.007 |
| 12 | 21843576 | rs11046164 | C | T | 2.89E-02 | 0.0046 |
| 12 | 45868623 | rs2408252 | C | T | -2.64E-02 | 0.0043 |
| 12 | 54429385 | rs12422600 | G | A | 2.40E-02 | 0.004 |
| 12 | 6691452 | rs7316626 | G | A | -3.22E-02 | 0.0054 |
| 12 | 117723613 | rs884847 | G | A | -2.93E-02 | 0.005 |
| 12 | 132544643 | rs11830241 | C | T | -4.11E-02 | 0.0072 |
| 12 | 106288445 | rs12825669 | G | A | 2.26E-02 | 0.0041 |
| 13 | 80705315 | rs11616380 | G | T | 7.64E-02 | 0.0043 |
| 13 | 26776999 | rs34584161 | G | A | -5.65E-02 | 0.0044 |

| | | | | | | |
|---|---|---|---|---|---|---|
| 13 | 91949562 | rs9515905 | G | A | -4.72E-02 | 0.0044 |
| 13 | 51094114 | rs9316500 | G | T | -4.01E-02 | 0.0041 |
| 13 | 23309382 | rs314879 | C | T | 3.64E-02 | 0.0046 |
| 13 | 41688401 | rs4397977 | G | A | -2.83E-02 | 0.0042 |
| 13 | 109946882 | rs9587811 | C | A | 2.48E-02 | 0.0038 |
| 13 | 54107583 | rs9568868 | G | T | -3.50E-02 | 0.0053 |
| 13 | 66204880 | rs9564268 | C | T | -2.43E-02 | 0.004 |
| 13 | 31017268 | rs12856169 | G | A | 3.64E-02 | 0.0061 |
| 13 | 50431987 | rs4942883 | C | A | 3.06E-02 | 0.0053 |
| 13 | 97176585 | rs61967710 | G | A | -5.54E-02 | 0.0099 |
| 14 | 79942647 | rs7156625 | G | A | -5.30E-02 | 0.0049 |
| 14 | 38804675 | rs7147483 | C | T | -3.52E-02 | 0.0041 |
| 14 | 33303540 | rs12883788 | C | T | -3.27E-02 | 0.004 |
| 14 | 101258584 | rs112324411 | C | T | 7.29E-02 | 0.0103 |
| 14 | 91963722 | rs8010382 | G | A | 2.66E-02 | 0.0038 |
| 14 | 69459229 | rs242105 | C | A | 3.90E-02 | 0.0059 |
| 14 | 77300863 | rs2056857 | C | T | 2.56E-02 | 0.0039 |
| 14 | 103376031 | rs4906272 | C | T | -2.99E-02 | 0.0046 |
| 14 | 61229411 | rs4902002 | G | A | 2.46E-02 | 0.0041 |
| 14 | 25947436 | rs11159347 | C | T | -2.48E-02 | 0.0041 |
| 14 | 47304091 | rs723355 | G | A | 2.37E-02 | 0.0039 |
| 14 | 58732748 | rs12892257 | G | A | 2.88E-02 | 0.005 |
| 14 | 30086481 | rs12433335 | C | T | -2.13E-02 | 0.0038 |

| | | | | | | |
|---|---|---|---|---|---|---|
| 14 | 103960026 | rs56365443 | G | A | -2.38E-02 | 0.0042 |
| 14 | 94039845 | rs11848361 | G | A | 4.78E-02 | 0.0085 |
| 15 | 77782335 | rs12910361 | G | A | 6.86E-02 | 0.004 |
| 15 | 90381278 | rs893617 | C | T | 5.37E-02 | 0.0041 |
| 15 | 91512067 | rs2290203 | G | A | -5.06E-02 | 0.0043 |
| 15 | 62391608 | rs7163757 | C | T | 4.30E-02 | 0.0038 |
| 15 | 75814388 | rs6495182 | C | T | 4.05E-02 | 0.0042 |
| 15 | 38828140 | rs8043085 | G | T | -4.07E-02 | 0.0042 |
| 15 | 40634717 | rs4923864 | G | A | 5.68E-02 | 0.0061 |
| 15 | 52587740 | rs149336329 | G | T | 8.69E-02 | 0.01 |
| 15 | 63871292 | rs7178762 | C | T | 3.23E-02 | 0.0041 |
| 15 | 74328576 | rs9479 | G | A | 2.71E-02 | 0.0037 |
| 15 | 57369850 | rs28490139 | G | A | 3.75E-02 | 0.0053 |
| 15 | 41801512 | rs2289739 | G | T | -3.64E-02 | 0.0055 |
| 15 | 93925327 | rs4777857 | G | A | 2.55E-02 | 0.0039 |
| 15 | 43895118 | rs2447198 | C | T | -3.40E-02 | 0.0055 |
| 15 | 39712286 | rs148106383 | C | T | -9.28E-02 | 0.0156 |
| 15 | 99276521 | rs59646751 | G | T | -2.41E-02 | 0.0041 |
| 15 | 84547222 | rs1812707 | C | T | -2.27E-02 | 0.0039 |
| 15 | 83461873 | rs36111056 | G | A | 3.03E-02 | 0.0053 |
| 15 | 49794020 | rs7169799 | C | T | 2.13E-02 | 0.0038 |
| 15 | 58676821 | rs11858759 | G | A | 2.36E-02 | 0.0043 |
| 16 | 53800954 | rs1421085 | C | T | 1.18E-01 | 0.004 |

| | | | | | | |
|---|---|---|---|---|---|---|
| 16 | 69666683 | rs244415 | G | A | 4.01E-02 | 0.0041 |
| 16 | 300388 | rs55857387 | C | T | -4.41E-02 | 0.0046 |
| 16 | 29958216 | rs8054556 | G | A | -3.17E-02 | 0.0039 |
| 16 | 3656482 | rs8061528 | C | T | -3.46E-02 | 0.0045 |
| 16 | 28897452 | rs8056890 | G | A | -2.95E-02 | 0.0042 |
| 16 | 85716463 | rs11646052 | G | A | 2.60E-02 | 0.0039 |
| 16 | 967241 | rs12918782 | G | A | 3.08E-02 | 0.0047 |
| 16 | 89630630 | rs12932337 | C | T | -3.40E-02 | 0.0052 |
| 16 | 88554480 | rs9937296 | C | T | 4.45E-02 | 0.007 |
| 16 | 87856424 | rs4384608 | C | T | 2.66E-02 | 0.0043 |
| 16 | 15153717 | rs9927842 | C | T | -3.07E-02 | 0.005 |
| 16 | 54387084 | rs2216063 | G | A | -2.96E-02 | 0.0054 |
| 16 | 70660243 | rs13330163 | G | A | -2.10E-02 | 0.0038 |
| 17 | 36099840 | rs11651755 | C | T | 6.63E-02 | 0.0038 |
| 17 | 3988451 | rs8071043 | C | T | 4.65E-02 | 0.0041 |
| 17 | 47060322 | rs35895680 | C | A | 4.99E-02 | 0.0047 |
| 17 | 40706273 | rs676387 | C | A | -4.16E-02 | 0.0041 |
| 17 | 65825248 | rs12603589 | C | T | 4.21E-02 | 0.0046 |
| 17 | 61565025 | rs4335 | G | A | 3.04E-02 | 0.0038 |
| 17 | 76772288 | rs7224711 | C | T | 3.07E-02 | 0.0038 |
| 17 | 29628549 | rs2040792 | C | A | 3.10E-02 | 0.0039 |
| 17 | 37746307 | rs11078916 | C | T | -3.26E-02 | 0.0042 |
| 17 | 62203059 | rs57767539 | G | A | -6.22E-02 | 0.0091 |

| | | | | | | |
|---|---|---|---|---|---|---|
| 17 | 41456413 | rs56799554 | G | A | 3.27E-02 | 0.0049 |
| 17 | 46178674 | rs3744347 | G | A | -3.01E-02 | 0.0046 |
| 17 | 70645032 | rs61736066 | G | A | 4.21E-02 | 0.0064 |
| 17 | 27570622 | rs9913225 | G | A | 2.62E-02 | 0.004 |
| 17 | 9787845 | rs17810376 | G | A | 2.79E-02 | 0.0047 |
| 17 | 78757626 | rs11150745 | G | A | -3.12E-02 | 0.0053 |
| 17 | 34862220 | rs1109442 | C | T | 2.18E-02 | 0.0037 |
| 17 | 481604 | rs11870735 | C | T | -3.14E-02 | 0.0054 |
| 17 | 21284910 | rs117642733 | C | T | -7.62E-02 | 0.0133 |
| 17 | 4854480 | rs366577 | C | T | 2.22E-02 | 0.004 |
| 18 | 60845884 | rs12454712 | C | T | -4.24E-02 | 0.0039 |
| 18 | 53050646 | rs72926932 | C | A | 7.33E-02 | 0.008 |
| 18 | 21083738 | rs303760 | C | T | -3.34E-02 | 0.0044 |
| 18 | 36746623 | rs7227272 | G | A | 4.08E-02 | 0.0056 |
| 18 | 74558999 | rs6565922 | C | T | -2.79E-02 | 0.0039 |
| 18 | 7070642 | rs7240767 | C | T | 3.40E-02 | 0.0051 |
| 18 | 63426979 | rs2032217 | G | A | -2.68E-02 | 0.004 |
| 18 | 56879827 | rs9319943 | C | T | -3.08E-02 | 0.0048 |
| 18 | 40066006 | rs410150 | C | T | 2.89E-02 | 0.0048 |
| 18 | 31582890 | rs17747955 | C | T | 2.24E-02 | 0.004 |
| 18 | 4845027 | rs9958640 | G | A | -2.36E-02 | 0.0042 |
| 19 | 46158417 | rs8107527 | G | A | -5.42E-02 | 0.004 |
| **19** | **19379549** | ***rs58542926** | **C** | **T** | **-7.21E-02** | **0.0072** |

| | | | | | | |
|---|---|---|---|---|---|---|
| 19 | 13010643 | rs9384 | G | T | 4.04E-02 | 0.0041 |
| 19 | 33896432 | rs4805881 | C | A | -3.79E-02 | 0.0039 |
| 19 | 7968168 | rs2115107 | G | A | -3.57E-02 | 0.0039 |
| 19 | 7240848 | rs75253922 | C | T | 4.02E-02 | 0.0054 |
| 19 | 47597102 | rs10408163 | C | T | 3.19E-02 | 0.0043 |
| 19 | 12505873 | rs7246440 | G | A | 2.99E-02 | 0.0043 |
| 19 | 31865946 | rs2867570 | G | A | 2.60E-02 | 0.0039 |
| 19 | 4967739 | rs12185519 | C | T | -3.60E-02 | 0.0054 |
| 19 | 18834514 | rs10404726 | C | T | 2.55E-02 | 0.0042 |
| 19 | 50016759 | rs142385484 | C | T | 3.36E-02 | 0.006 |
| 19 | 1646712 | rs4807125 | C | T | -3.26E-02 | 0.0059 |
| 20 | 42994812 | rs12625671 | C | T | 6.03E-02 | 0.0055 |
| 20 | 45594711 | rs6066138 | G | A | 4.20E-02 | 0.0047 |
| 20 | 51033681 | rs4809906 | G | A | 3.45E-02 | 0.004 |
| **20** | **32675727** | ***rs6059662** | **G** | **A** | **3.69E-02** | **0.0043** |
| 20 | 57396495 | rs4810145 | C | T | 3.03E-02 | 0.0038 |
| 20 | 62450664 | rs6011155 | C | T | -2.84E-02 | 0.0039 |
| 20 | 50155386 | rs6021276 | C | T | -2.83E-02 | 0.004 |
| 20 | 42230695 | rs6073143 | C | T | 3.04E-02 | 0.0044 |
| 20 | 51620857 | rs2252115 | G | A | -2.37E-02 | 0.0039 |
| 20 | 22428284 | rs7274134 | C | T | 2.47E-02 | 0.0041 |
| 20 | 39832628 | rs17265513 | C | T | 3.27E-02 | 0.0055 |
| 20 | 2100095 | rs6137042 | G | A | 2.63E-02 | 0.0047 |

| | | | | | | |
|---|---|---|---|---|---|---|
| 22 | 44324855 | rs3747207 | G | A | -4.12E-02 | 0.0044 |
| 22 | 50356850 | rs1801645 | C | T | 3.74E-02 | 0.0045 |
| 22 | 29369398 | rs5762925 | C | A | 2.67E-02 | 0.0039 |
| 22 | 41593581 | rs11913442 | C | T | -2.45E-02 | 0.004 |
| 22 | 35705359 | rs138771 | G | A | -2.59E-02 | 0.0044 |
| 22 | 32203334 | rs75307421 | G | A | -7.37E-02 | 0.0132 |
